# SARS-CoV-2 serological testing using electrochemiluminescence reveals a rapid onset of seroconversion in severe COVID-19 patients

**DOI:** 10.1101/2020.06.28.20141838

**Authors:** A Munitz, L Edry-Botzer, M Itan, R Tur-Kaspa, D Dicker, D Markovitch, M Goren, M Mor, S Lev, T Gottesman, K Muhsen, D Cohen, M Stein, U Qimron, N Freund, Y Wine, M Gerlic

## Abstract

Despite ongoing efforts to characterize the host response toward SARS-CoV-2, a major gap in our knowledge still exists regarding the magnitude and duration of the humoral response. We report the development of a rapid, highly specific and sensitive electrochemiluminescent assay for detecting IgM, IgA, and IgG antibodies toward two distinct SARS-CoV-2 antigens namely, the receptor binding domain (RBD) and the nuclear protein (NP). Whereas IgM antibodies toward RBD were detected at early stages of the disease, IgM antibodies against NP did not develop. Analysis of the antibody response in mild versus moderate/severe patients revealed a rapid onset of IgG and IgA antibodies, specifically in moderate/severe patients. Finally, we observed a marked reduction in IgM/IgA antibodies and to lesser extent, IgG, over time. We provide a comprehensive analysis of the human antibody response, and has major implications on our understanding and monitoring of SARS-CoV-2 infections, as well as finding effective vaccines.

**One Sentence Summary:** Using a newly developed assay to detect anti-SARS-Cov-2 IgM, IgG and IgA antibodies we reveal a rapid onset of IgG and IgA antibodies towards distinct viral antigens, specifically in moderate/severe COVID-19 patients,

## Introduction

The eruption of the COVID-19 pandemic, caused by the newly discovered SARS-CoV-2 virus, has had a profound impact on human life on a global scale^1,2^. COVID-19 has affected and still affects millions of people worldwide, resulting in high mortality and morbidity rates as well as high health care costs and difficulties in treatment^3^. Furthermore, unprecedented government interventions indirectly caused significant morbidity and mortality. This is exemplified by the intense engagement of most health facilities with COVID-19; consequently, they were unavailable to patients suffering from other diseases and conditions^4^. In addition, the overwhelming economic burden that COVID-19 imposes on most countries is expected to result in the loss of numerous additional lives, including health care workers, along with extensive long-term damage^5^.

Detection of infected individuals is typically carried out by using quantitative PCR (qPCR) analysis, which amplifies viral genes. Although this method is an excellent tool for surveillance of viral spread, it has major drawbacks, including decreased accuracy when swabs are taken 5 days after the symptoms appear (∼70%)^6^. Furthermore, it is expensive and does not provide substantial data on the immunity of a given individual in the population. Thus, although excellent tools exist for the diagnosis of viral load and the diagnosis of infected individuals, a major gap still exists in understanding and effectively responding to the host. Specifically, the kinetics of the humoral immune response following SARS-CoV-2 infection and the association between the emergence of different antibody subsets and the disease severity are largely unknown^7^.

The main hurdle in generating such knowledge is the development of reliable diagnostic tools, which will enable sensitive and accurate serological testing. However, the presence of multiple a-symptomatic individuals and the fact that it remains unclear whether antibodies are generated with the onset of symptoms strengthen the need for kinetic analysis of the host response using rapid and accurate serological assays. Owing to the relative accessibility of blood sampling (in comparison with nasopharyngeal swabs) and the relatively high stability of antibodies, the development of such assays will provide excellent tools for epidemiological studies by screening of populations and better understanding the spread of SARS-CoV-2. In addition, reliable serological tests can provide critical clinical information regarding the course of the disease and the host response^8^. Finally, since mucosal tissues, such as the respiratory tract, are affected in COVID-19, it is extremely important to monitor the differential expression of IgM, IgG, and IgA antibodies and to correlate them with the clinical outcomes^9^.

The invaluable insights that can be achieved by serological tests prompted us to develop an accurate and sensitive assay that can detect the three main antibody classes, IgM, IgG, and IgA, using electrochemiluminescence. Our assay was validated using 96 samples from different COVID-19 patients and 195 serum samples that were obtained before November 2019. In subsequent kinetic analyses, we determined the seropositivity of IgM, IgG, and IgA antibodies toward the receptor binding domain of the spike protein (RBD) as well as the N-protein of SARS-CoV-2 (NP). Our data revealed that IgM is less generated against NP antigen, whereas high titers were detected against RBD antigen. Finally, we compared the clinical parameters with the antibody response in mild versus moderate/severe patients. Our analyses identified a rapid onset of IgG and IgA antibodies specifically in moderate/severe patients in comparison with the mild ones. Collectively, this study provides a comprehensive analysis of the human antibody response, and has major implications on our understanding and monitoring of SARS-CoV-2.

## Results

### Development of SARS-CoV-2 electrochemiluminescence-based ELISA

In attempting to develop a serological assay for detecting antibodies toward SARS-CoV-2 antigens, we sought to calibrate an electroluminescence-based ELISA test according to the following criteria: First, the assay should be accurate and should display >95% sensitivity and >97% specificity. Second, the assay should be informative and potentially enable multiplexing of several antibody classes toward different SARS-CoV-2 antigens. Finally, the assay should be robust so that it can be upscaled in regular hospitals and/or community health laboratories to enable the testing of multiple individuals simultaneously.

To this end, we hypothesized that an electrochemiluminescent-based assay would be an ideal platform^10,11^. Electrochemiluminescence is a type of luminescence that is produced during electrochemical reactions in solution. Such assays provide a high dynamic range and are fully quantitative. Furthermore, by placing high binding carbon electrodes at the bottom of multi-spot microplates, such assays allow easy attachment of multiple biological reagents such as SARS-CoV-2 antigens and potentially enable high-throughput multiplexing.

First, we performed a side-by-side comparison between a standard enzymatic (i.e., horseradish peroxidase-based) ELISA test (termed HRP) and an electrochemiluminescence-based ELISA test (termed TauMed). The standard ELISA test could detect IgG and IgM antibodies against the SARS-CoV-2 RBD (Figure 1A-B, respectively). However, the HRP-based ELISA test was non-linear in the lower serum dilutions, both in IgM and IgG (Figure 1A-B). In contrast, the electrochemiluminescence-based test (TauMed) displayed a linear titration, which was observed in the diluted COVID-19-positive serum. (Figure 1C-D). Furthermore, a dose-dependent signal-to-noise ratio, which was observed in the TauMed electrochemilumines-cence test, was observed and compared with the regular HRP test (Figure 1E-F). Finally, the TauMed electrochemiluminescence test showed a higher sensitivity (∼100-fold for IgG and ∼30-fold for IgM), in comparison with HRP (∼10-fold for both IgG and IgM).

**Figure 1.**
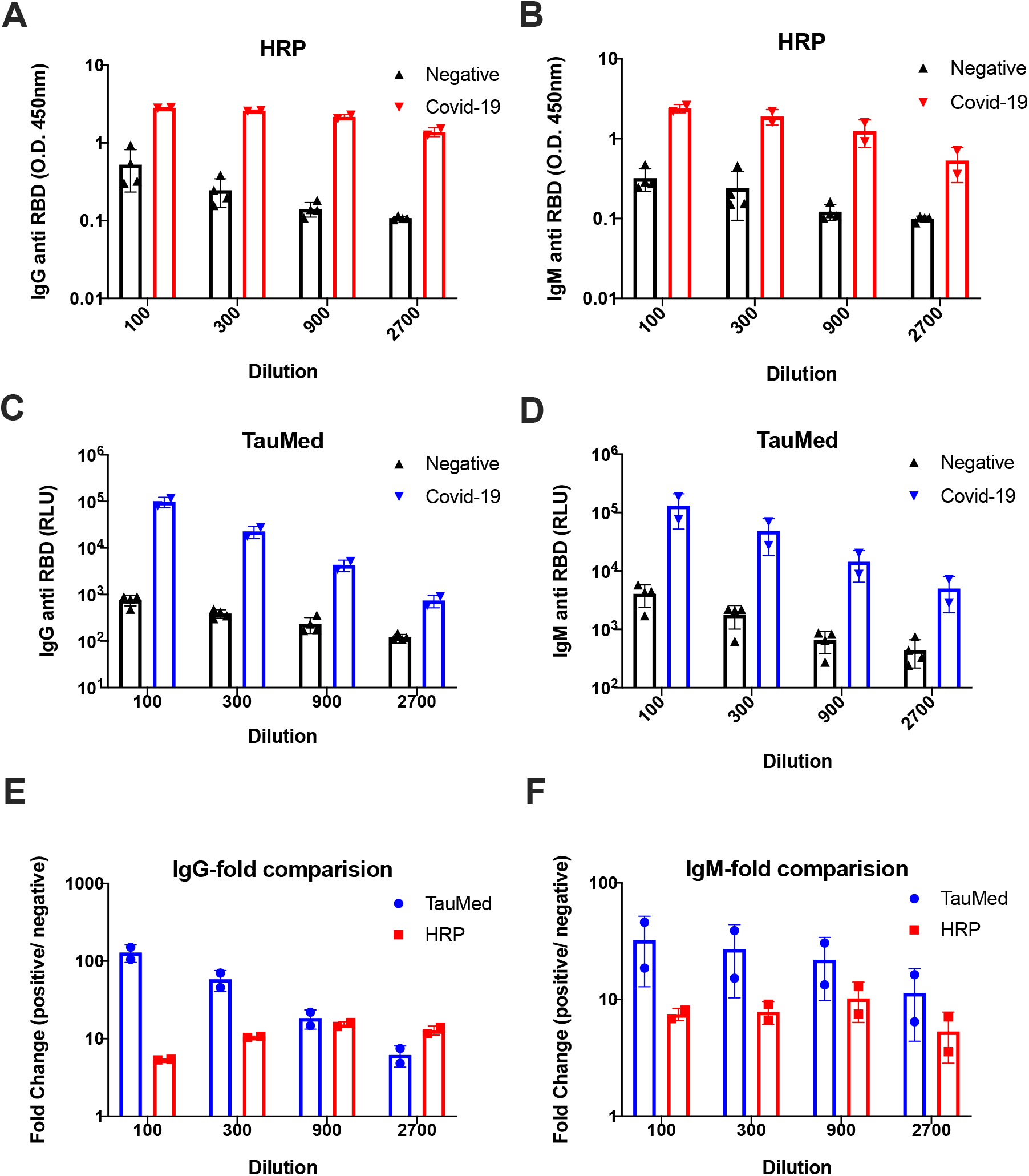
Comparison of SARS-CoV-2 HRP-based ELISA to electrochemiluminescence-based ELISA. The presence of anti-RBD IgG and IgM antibodies was determined in a side-by-side comparison using HRP-based ELISA (HRP) (A, B) and electrochemiluminescence-based ELISA (TauMed) (C, D). The fold changes in the values obtained from individual positive samples over the average negative samples were calculated (E, F).

### Validation of SARS-CoV-2 electrochemiluminescence-based ELISA

To validate our assay and determine the cutoff range for assay specificity and sensitivity, we obtained sera from multiple COVID-19 patients (the patient characteristics are summarized in Table 1), multiple COVID-19 recovered patients, as well as sera from patients that were not exposed to SARS-CoV-2 (e.g., serum that was obtained before November 2019). A significant increase in electrochemiluminescence was observed in our patient cohort, demonstrating the presence of anti-RBD IgG, IgM, and IgA antibodies (Figure 2A-C). Using ROC analysis of sera from >14 days post symptoms (DPS) from COVID-19 patients and recovered individuals (n=68) and all of the negative samples (n=195 for IgG and IgM and n=97 for IgA), we determined a cutoff value of ∼95% and ∼98% specificity (Wilson/Brown 95% CI) and the equivalent sensitivity for these cutoffs for all three antibodies (Fig. 2D-F and Table 2). Since individual patients may test positive to one or two out of the three antibody classes, we further analyzed our data using a combined IgG, IgM, and IgA strategy. In this analysis, positivity toward COVID-19 seroconversion was determined by testing positive (using the ∼98% specificity) for only one out of three specific RBD antibody classes. This new combined analysis resulted in 94.9% specificity and increased the sensitivity from ∼78-91% to 100% for the >14 DPS COVID-19 patients and recovered individuals. Using individual and combined approaches, we analyzed all patient samples that were also obtained at earlier time points and that had post symptoms (i.e., <14 DPS). This approach could detect COVID-19 seropositive patients even in the early stages of the disease (Figure 2G-H and Table 3) In fact, the sensitivity increased from ∼46-61% for a specific antibody class to 84.6% using the combined strategy at ≤7 DPS and from ∼40-73.3% to 80% in the second week of post symptoms. In total, regardless of DPS, the sensitivity increased from ∼75-79% to 94.8%.

**Table 1.** Patient characteristics

| Disease severity | Gender | N= | Age | DPS | Lymphocyte count | CRP |
| --- | --- | --- | --- | --- | --- | --- |
| Mild (N=29) | Female | 13 | 66.58 ± 19.07 | 18.08 ± 18.64 | 1.75 ± 0.63 | 0.74 ± 0.66 |
|  | Male | 16 | 65.41 ± 14.68 | 29.75 ± 23.73 | 1.4 ± 0.63 | 5.66 ± 4.78 |
| Moderate /Severe (N=28) | Female | 5 | 76.20 ± 4.71 | 16.00 ± 14.68 | 2.70 ± 4.36 | 6.96 ± 5.37 |
|  | Male | 23 | 67.16 ± 15.36 | 21.22 ± 16.14 | 1.64 ± 2.10 | 5.10 ± 7.76 |

**Table 2.**
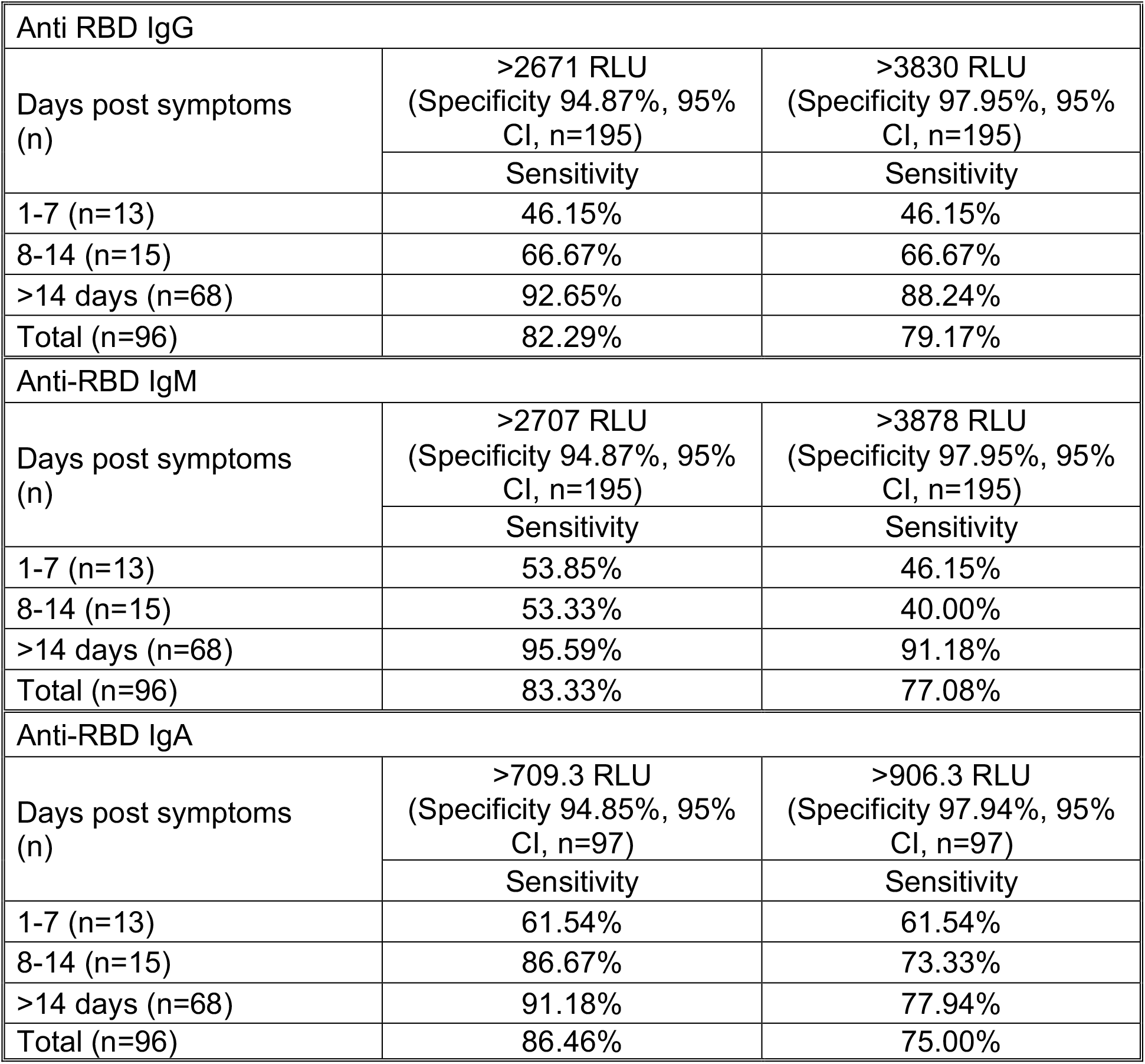
anti RBD antigen

**Table 3.**
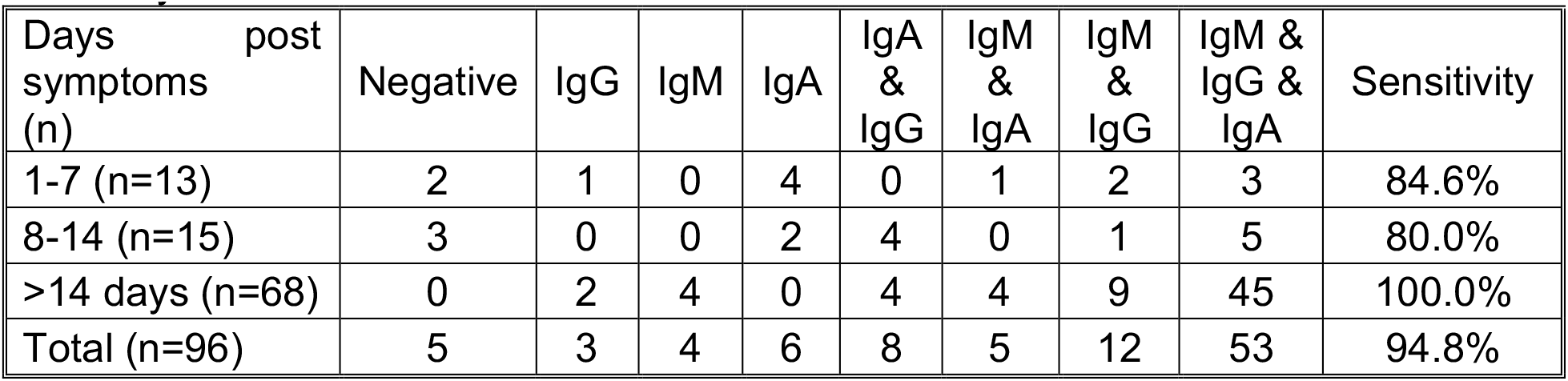
Anti-RBD combined analysis using 98% specificity for each individual antibody

| Days post<br>symptoms<br>(n) | Negative | IgG | IgM | IgA | IgA<br>&<br>IgG | IgM<br>&<br>IgA | IgM<br>&<br>IgG | IgM &<br>IgG &<br>IgA | Sensitivity |
| --- | --- | --- | --- | --- | --- | --- | --- | --- | --- |
| 1-7 (n=13) | 2 | 1 | 0 | 4 | 0 | 1 | 2 | 3 | 84.6% |
| 8-14 (n=15) | 3 | 0 | 0 | 2 | 4 | 0 | 1 | 5 | 80.0% |
| >14 days (n=68) | 0 | 2 | 4 | 0 | 4 | 4 | 9 | 45 | 100.0% |
| Total (n=96) | 5 | 3 | 4 | 6 | 8 | 5 | 12 | 53 | 94.8% |

**Figure 2.**
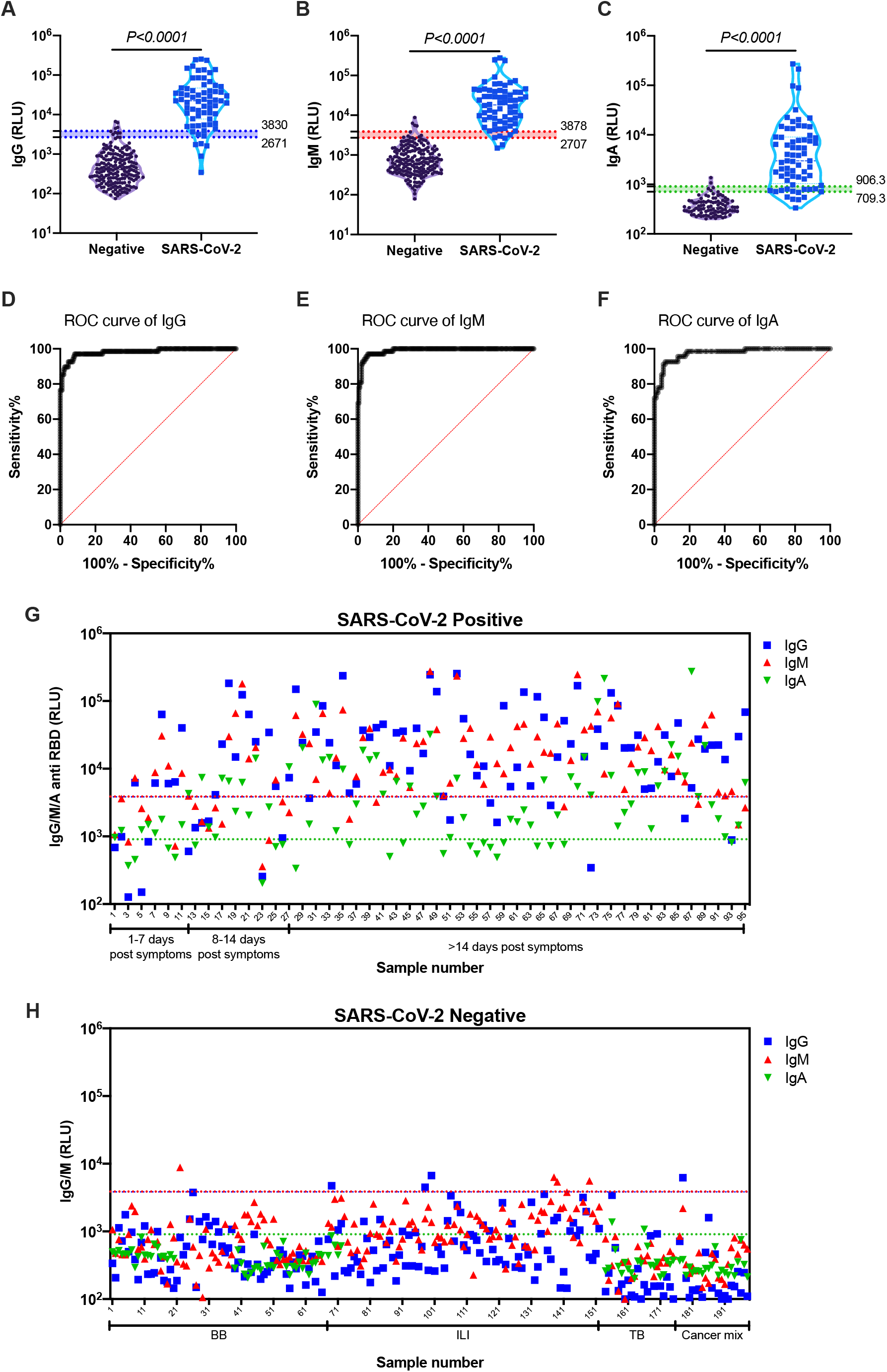
Validation of anti-SARS-CoV-2-RBD antibodies using electrochemiluminescence ELISA. Peripheral blood was collected from the peripheral blood of COVID-19 patients and recovered patients (n=96). Negative samples were obtained from true SARS-CoV-2 negative patients (i.e., prior to the SARS-CoV-2 pandemic) (n=197). Plasma was obtained, diluted 1:50, and added to a 96-well plate precoated with SARS-CoV-2 RBD antigen. IgG (A), IgM (B), and IgA (C) levels as well as ROC analysis (D-F) are shown. Individual IgG (blue), IgM (red), and IgA (green) levels of each SARS-CoV-2 positive (G) and a negative sample (H) are shown. Data were calculated using GraphPad Prism 8; the dotted line represents the calculated cutoff value discriminating between positive and negative samples. (A-C) A nonparametric Mann-Whitney t-test was performed. P values are shown.

The specific immune response toward distinct viral antigens may result in different kinetics of the humoral antibody response^12^. Thus, we aimed to determine the presence of antibodies toward an intra-viral protein/antigen, which may be presented to the immune system at later times post-infection. This is in addition to antibodies directed against the RBD domain, which is part of the viral surface protein spike and thus is more likely to be presented to the immune system at earlier times post-infection. To this end, we used the nucleocapsid protein (NP) as an additional antigen. A significant increase in anti-NP IgG and IgA antibodies (Figure 3A, 3C) but not IgM antibodies (Figure 3B) was observed in our patient cohort. Using ROC analysis of the >14 DPS COVID-19 patients (n=31) and negative samples (n=90), we determined a cutoff value for achieving ∼95% and ∼98% specificity (Wilson/Brown 95% CI) and the equivalent sensitivity for all three antibodies (Figure 3D-F and Table 4). Furthermore, we employed our combined IgG, IgM, and IgA analysis strategy (Figure 3G-H and Table 5). Since the NP antigen appears to elicit an antibody response that is primarily IgG, this combined analysis strategy was less efficient than what we obtained when analyzing anti-RBD antibody responses.

**Table 4.** Anti-NP antigen

| Anti NP IgG |  |  |
| --- | --- | --- |
| Days post symptoms (n) | >1245 RLU<br>(Specificity 95.56%, 95% CI, n=90) | >1995 RLU<br>(Specificity 97.78%, 95% CI, n=90) |
|  | Sensitivity | Sensitivity |
| 1-7 (n=13) | 53.85% | 53.85% |
| 8-14 (n=13) | 61.54% | 53.85% |
| >14 days (n=31) | 96.77% | 96.77% |
| Total (n=57) | 78.95% | 77.19% |
| Anti-NP IgM |  |  |
| Days post symptoms (n) | >5982 RLU<br>(Specificity 95.56%, 95% CI, n=90) | >7466 RLU<br>(Specificity 97.78%, 95% CI, n=90) |
|  | Sensitivity | Sensitivity |
| 1-7 (n=13) | 0% | 0% |
| 8-14 (n=13) | 30.77% | 30.77% |
| >14 days (n=31) | 32.26% | 32.26% |
| Total (n=57) | 24.56% | 24.56% |
| Anti-NP IgA |  |  |
| Days post symptoms (n) | >1891 RLU<br>(Specificity 95.56%, 95% CI, n=90) | >4500 RLU<br>(Specificity 97.78%, 95% CI, n=90) |
|  | Sensitivity | Sensitivity |
| 1-7 (n=13) | 15.38% | 0% |
| 8-14 (n=13) | 53.85% | 53.85% |
| >14 days (n=31) | 87.10% | 54.84% |
| Total (n=57) | 63.16% | 42.11% |

**Table 5.** Anti-NP combined analysis using 98% specificity for each individual antibody

| Days post<br>symptoms<br>(n) | Negative | IgG | IgM | IgA | IgA<br>&<br>IgG | IgM<br>&<br>IgA | IgM<br>&<br>IgG | IgM &<br>IgG &<br>IgA | Sensitivity |
| --- | --- | --- | --- | --- | --- | --- | --- | --- | --- |
| 1-7 (n=13) | 6 | 7 | 0 | 0 | 0 | 0 | 0 | 0 | 53.8% |
| 8-14 (n=13) | 5 | 1 | 0 | 1 | 2 | 0 | 0 | 4 | 61.5% |
| >14 days (n=31) | 1 | 11 | 0 | 0 | 9 | 0 | 2 | 8 | 96.8% |
| Total (n=57) | 12 | 19 | 0 | 1 | 11 | 0 | 2 | 12 | 78.9% |

**Figure 3.**
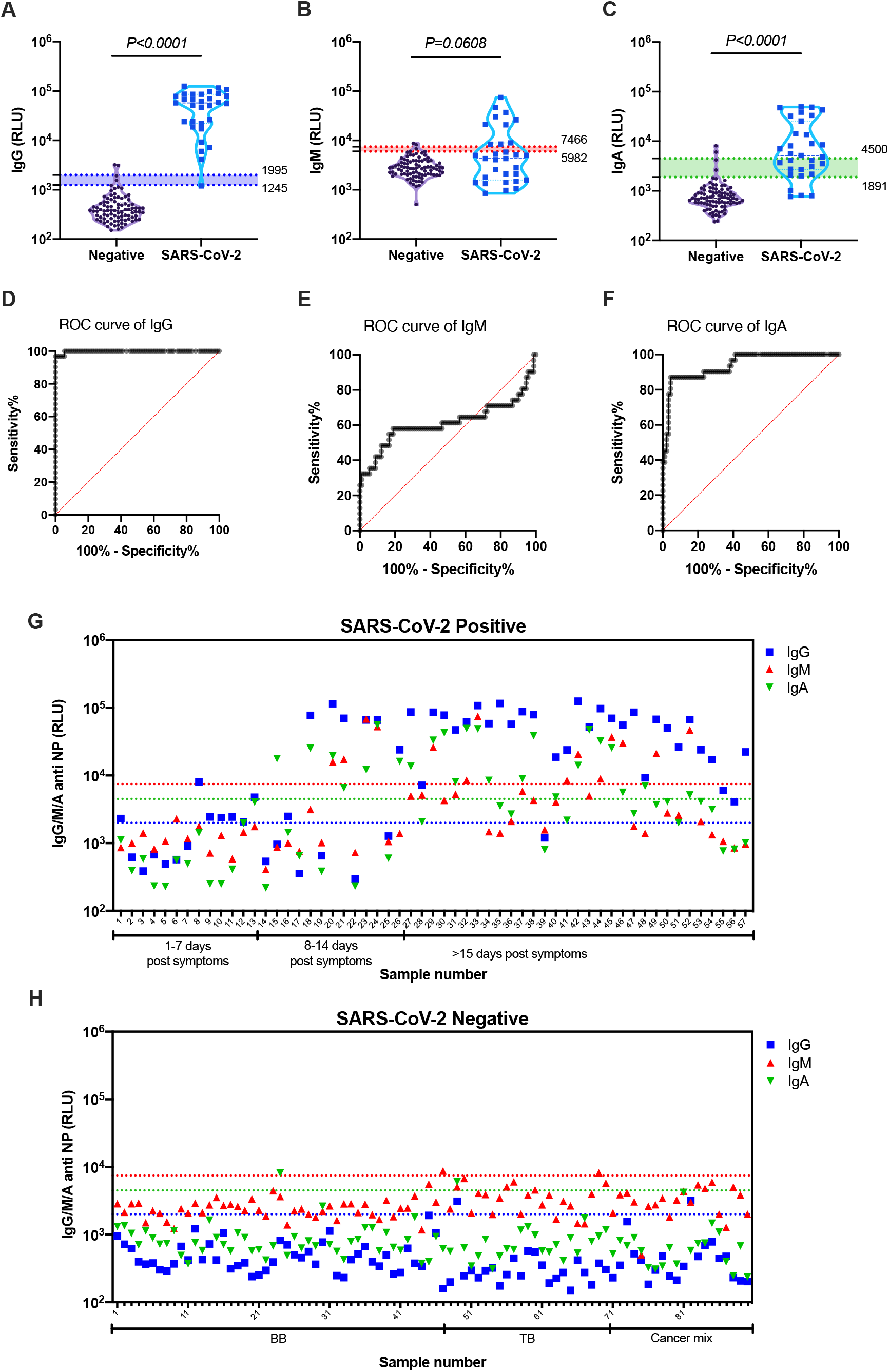
Validation of anti-SARS-CoV-2-NP antibodies using electrochemiluminescence ELISA. Peripheral blood was collected from the peripheral blood of COVID-19 and recovered patients (n=57). Negative samples were obtained from true SARS-CoV-2 negative patients (i.e., prior to the SARS-CoV-2 pandemic) (n=90). Plasma was obtained, diluted 1:50, and added to a 96-well plate precoated with SARS-CoV-2 NP antigen. IgG (A), IgM (B), and IgA (C) levels as well as ROC analysis (D-F) are shown. Individual IgG (blue), IgM (red), and IgA (green) levels of each SARS-CoV-2 positive (G) and negative sample (H) are shown. Data were calculated using GraphPad Prism 8; the dotted line represents the calculated cutoff value discriminating between positive and negative samples. (A-C) A nonparametric Mann-Whitney t-test was performed. P values are shown.

### SARS-CoV-2 RBD antigen as a serological marker shows superior results to NP antigen

Next, we compared the kinetics of the host antibody response in our patient population. To this end, we divided the patients into four groups: 1-7 DPS, 8-14 DPS, 15-28 DPS, and >29 DPS. Notably, the latter group consisted of either patients with active disease or recovered individuals. As shown in Figure 4, all antibody classes against SARS-CoV-2 RBD antigen developed rapidly, and were readily detected even in the patient groups that were in their first week post symptoms. In contrast, IgG and IgA antibodies against SARS-CoV-2 NP antigens develop much slower. In agreement with our hypothesis regarding the time of antigen exposure, IgM anti-NP antibodies did not develop within the first two weeks post symptoms. On the other hand, IgG antibodies against NP reached a peak similar to that of anti-RBD, showing a high specificity after two weeks.

**Figure 4.**
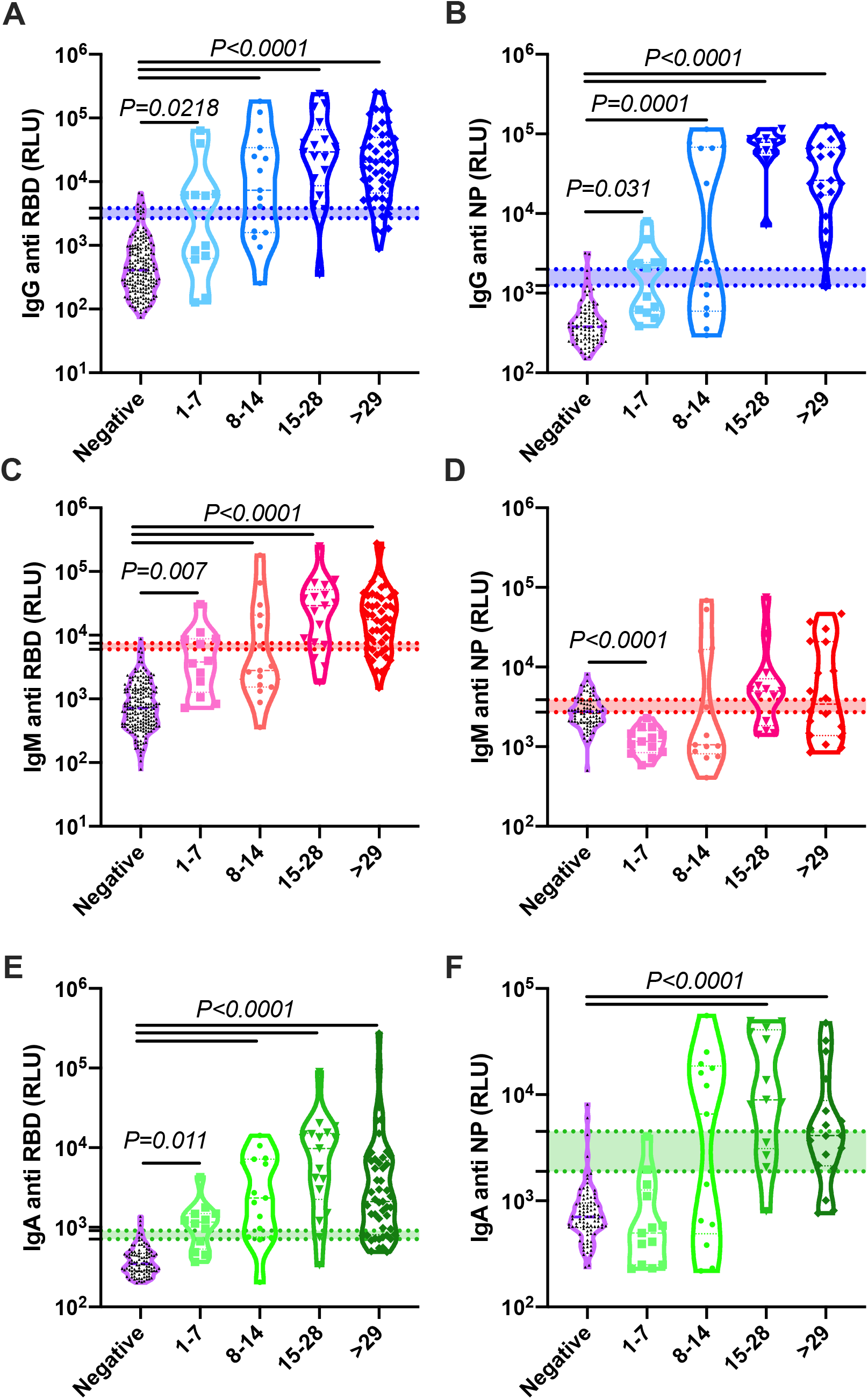
Comparison of the development anti-SARS-CoV-2 RBD and NP antibodies. Peripheral blood was collected from COVID-19 and recovered patients. Negative samples were obtained from true SARS-CoV-2 negative patients (i.e., prior to the SARS-CoV-2 pandemic). Plasma was obtained, diluted 1:50, and added to a 96-well plate precoated with SARS-CoV-2 RBD (A, C, and E) or NP (B, D, and F) antigens. IgG (A, B), IgM (C, D), and IgA (E, F) levels are shown. Data were calculated using GraphPad Prism 8; the dotted line represents the calculated cutoff values (95% and 98% sensitivity) discriminating between positive and negative samples. Statistical analysis was performed using a Nonparametric Kruskal-Wells test for multiple comparisons.

### Rapid onset of antibodies in moderate/severe patients in comparison to mild ones

To assess whether the antibody response during SARS-CoV-2 infection correlated with any clinical parameters, we divided our patient cohort into two groups consisting of mild and moderate/severe patients (see Table 1). A pooled analysis of all the sera antibody titers, independent of disease severity, revealed that antibodies (i.e., IgG, IgM, and IgA) against RBD and NP peaked between 15 and 28 DPS. IgM against NP as well as IgA against both antigens appeared to peak slightly later and maximal antibody titers were observed at 29-42 DPS (Figure 5A, B). These data are in agreement with the data presented in Figure 4, which presents recovered patients with no data on their disease severity. Notably, although still detectable, all antibody classes appear to start to decrease at 42 DPS, with the exception of IgG anti-RBD (Figure 5A). These data were further observed by analyses of individual patients, which were at several time points (Figure 5C-H).

**Figure 5.**
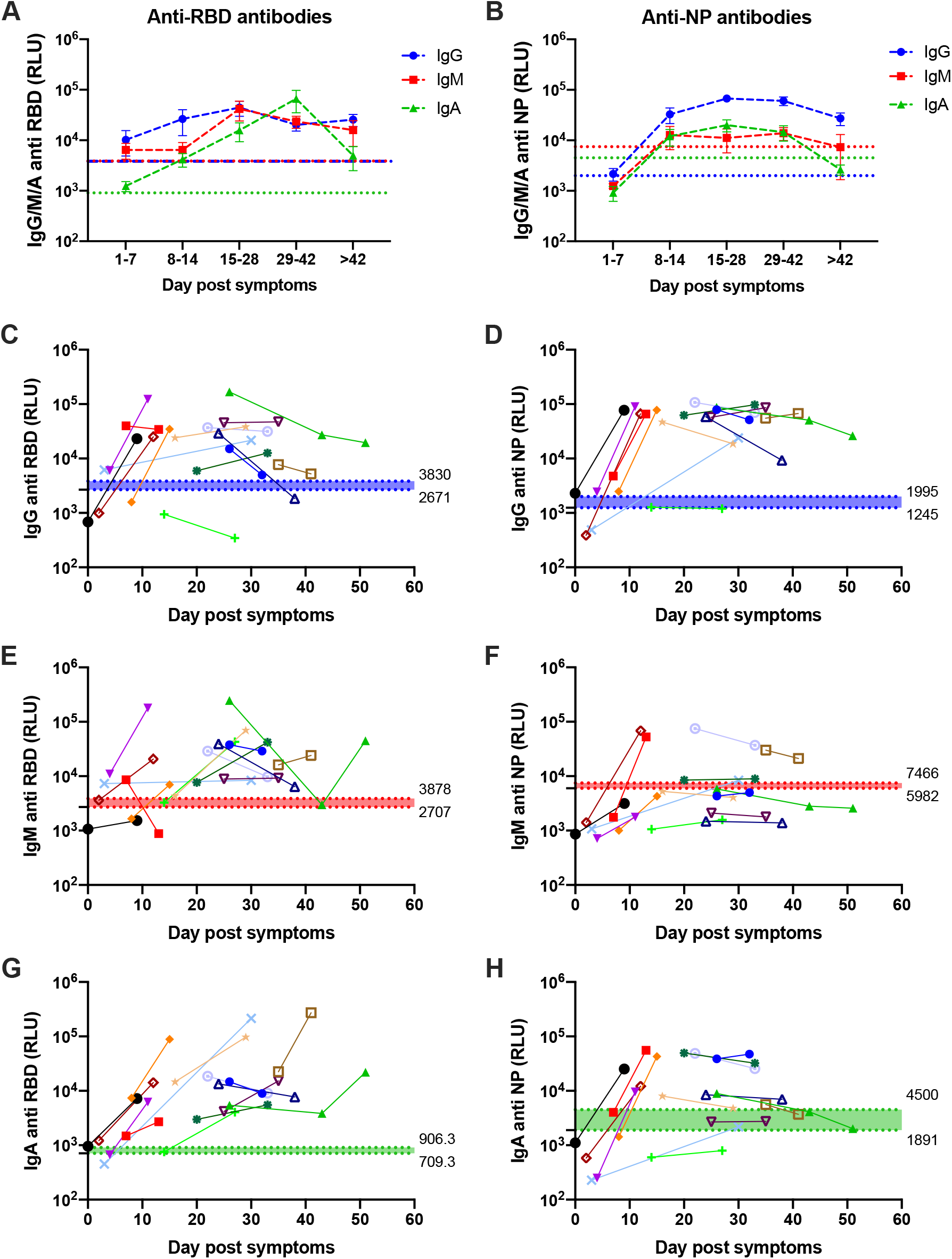
SARS-CoV-2 antibodies’ response kinetics. Peripheral blood was collected from COVID-19 patients. Plasma was obtained, diluted 1:50, and added to a 96-well plate precoated with SARS-CoV-2 RBD (A, C, E, and G) or NP (B, D, F, and H) antigens. A, B Kinetics of all samples; average±SEM. C-H Kinetics of individual patient’s antibody response. Data were calculated using GraphPad Prism 8; the dotted line represents the calculated cutoff values (98% sensitivity for A, B, and 95% and 98% sensitivity for C-H) discriminating between positive and negative samples.

Next, the readout for each antibody class was assessed in each patient group (see Table 1) separately (Figure 6). This analysis revealed that the onset of the antibody response was rapid in moderate/severe patients in comparison with mild patients. In fact, a rapid onset of IgG and IgA antibodies against RBD and NP was detected in moderate/severe patients as well as IgM antibodies against RBD in comparison with mild patients (Figure 6). Despite the slower kinetic pattern, all patients, regardless of their disease severity, could develop similar levels of antibodies. Although not statistically significant, an obvious reduction trend in IgM and IgA was observed in all the late stages of the disease (i.e., >42 DPS) in both populations of patients, whereas only a slight reduction was observed in IgG antibodies

**Figure 6.**
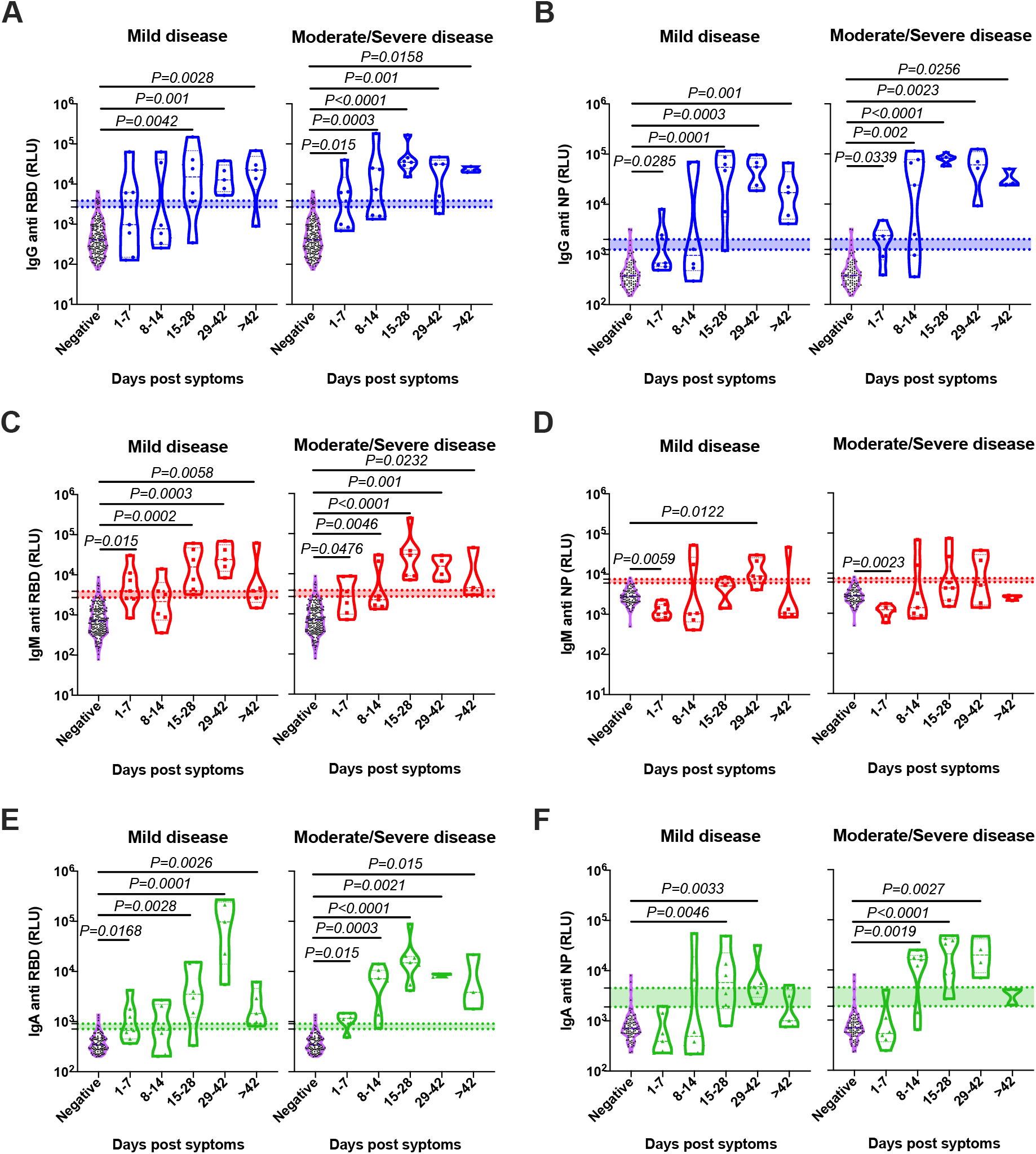
Correlation of the antibodies’ response to the disease severity. Peripheral blood was collected from COVID-19 patients. Negative samples were obtained from true SARS-CoV-2 negative patients (i.e., prior to the SARS-CoV-2 pandemic). Plasma was obtained, diluted 1:50, and added to a 96-well plate precoated with SARS-CoV-2 RBD (A, C, and E) or NP (B, D, and F) antigens. Patients’ antibody results were grouped according to their disease severity and graphed against DPS. Data were calculated using GraphPad Prism 8; the dotted line represents the calculated cutoff values (95% and 98% sensitivity) discriminating between positive and negative samples. Statistical analysis was performed using a Nonparametric Kruskal-Wells test for multiple comparisons against negative samples. Significant P values are shown.

### Antibody kinetics and its association with gender and clinical parameters

Previous data suggested that males are more susceptible to develop severe COVID-19 disease^13^. In support of these data, the male-to-female ratio in the mild COVID-19 patient cohort was 0.81, whereas in the moderate/severe patient population it was 0.22, demonstrating the predominance of males over females in moderate/severe patients (Table 1). Given this gender difference in our patient cohorts, we aimed to determine whether this may bias our data toward the rapid onset of antibodies in moderate/severe patients in comparison to mild ones (Figure 6). Thus, we assessed the onset of antibodies toward RBD and NP in males vs. females in each patient cohort. No significant differences were observed between the different genders (Figure 7). In addition, no significant correlations were observed between any antibody response and the levels of CRP and/or lymphocyte cell counts (Supplemental Figure 1).

**Figure 7.**
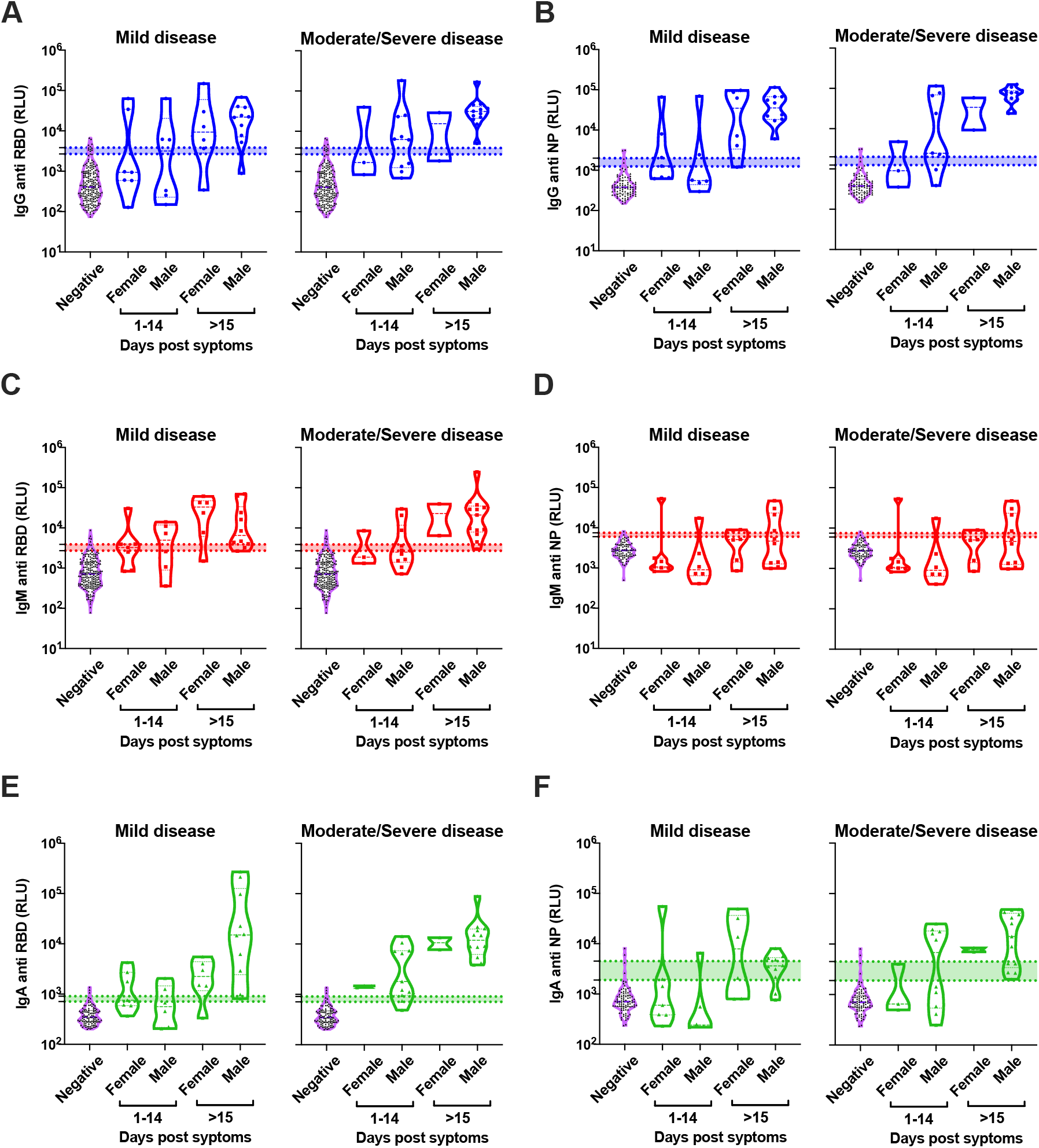
No correlation of antibodies’ response to patient gender. Peripheral blood was collected from COVID-19 patients. Negative samples were obtained from true SARS-CoV-2 negative patients (i.e., prior to the SARS-CoV-2 pandemic). Plasma was obtained, diluted 1:50, and added to a 96-well plate precoated with SARS-CoV-2 RBD (A, C, and E) or NP (B, D, and F) antigens. Patients’ antibody results were grouped according to their gender and the disease severity and graphed against DPS (1-14 and >15). Data were calculated using GraphPad Prism 8; the dotted line represents the calculated cutoff values (95% and 98% sensitivity) discriminating between positive and negative samples. Statistical analysis was performed using a Nonparametric Kruskal-Wells test for multiple comparisons against between female and male groups. No significant difference was found between genders.

## Discussion

Herein we describe a rapid, accurate, and robust serological method to detect seroconversion upon SARS-CoV-2 infection. Our method is based on the reactivity of the major classes of antibodies, namely, IgG, IgM, and IgA toward the immunogenic RBD and NP proteins of the virus. This method has several advantages over the standard ELISA procedures. For example, many standard ELISA tests, which are used for diagnostic serological testing, rely on enzymatic activity for the end point detection of the antibodies. This enzymatic activity introduces several limitations. Since the enzymatic reaction is time dependent, many ELISA tests require the use of a standard curve in order to detect the sample in the linear range of the assay. In electrochemiluminescence-based assays, such as the one we have developed, each individual sample in the plate is electronically excited and emits light, which is recorded immediately. This enables the assay to have a wide dynamic range that exceeds that of the standard ELISA tests. An additional advantage of the tests we developed is the high positive-to-negative ratio. Using a standard ELISA test, we could achieve a ∼10-fold induction, whereas using our assay we reached ∼100-fold. An additional advantage of the platform that we used is the ability to assess all three major antibody classes using multiplexing. This is extremely important in two different aspects. Regarding the diagnostic aspect, this allows us to increase the sensitivity of the assay by cross-analyzing the formation of different antibodies in each individual. Indeed, although many patients developed all three antibodies toward the viral RBD, several patients could generate only a single antibody class (e.g., they were positive for IgM but not for IgA or IgG). Such patients would be perceived as patients that did not develop antibodies if they were assessed by assays that enable the detection of 1 or even 2 antibodies. Strengthening this notion, our combined analysis strategy could increase the assay’s sensitivity by nearly 100%. This is noteworthy, since all of our analyses were conducted using a threshold that corresponds to ∼98% specificity to each individual antibody.

To better define the host antibody response toward different viral antigens, we compared the onset of antibody generation toward the SARS-CoV-2 antigens, RBD and NP. Our data indicate marked differences in the generation of an IgM response between these two distinct antigens. Whereas IgM antibodies toward RBD were readily detected even at an early stage post-symptoms, IgM antibodies against NP did not develop. This is most likely explained by the kinetics of viral entry and replication in mucosal epithelial cells^9^. Initial exposure of the immune system is probably initiated by external antigens (even in a low viral load), whereas only later on, when the viral load increases and perhaps immune-mediated epithelial cell and/or viral death occurs, internal antigens such as NP are exposed. The finding that different viral antigens elicit differential kinetics in terms of the antibody’s response is important, since multiple serological assays are currently being developed and each assay targets different antigens. It will be important to limit the conclusions of each study to the specific antigen that was examined. This may also be an underlying difference between different serological testing method results, where, for example, an individual generated antibodies toward RBD but not NP^14–16^. Directly related, the technological basis of our assay allows each well to be coated within a given 96-well plate with several antigens (up to 10 different antigens per well). This will enable rapid multiplexing of differential antibody responses toward several viral antigens within a given sample and will enable one to obtain a comprehensive understanding of the host response toward SARS-CoV-2.

In agreement with a previous publication^17^, our analysis of the antibody response in mild vs. moderate/severe patients revealed that moderate/severe patients generated a relatively rapid antibody response, especially toward RBD. Although our study bears the limitation that we do not know the time of infection and that the reference point for our analyses is the time post symptoms, these data clearly indicate that the disease severity is not due to a lack of antibody response in moderate/severe patients. In fact, our data corroborate previous publications describing increased immune cell activation and subsequently an increased pro-inflammatory response of the host in severe patients^18^. Thus, we believe that the rapid production of antibodies in moderate/severe individuals reflects this phenomenon. Unfortunately, we could not correlate the level of antibody response with the viral load of a given individual since we could not obtain the quantitative PCR Ct values of each patient. Future studies are required to establish such correlations.

Several studies raised the hypothesis that moderate/severe COVID-19 patients may elicit a harmful antibody response^19^. In this scenario, part of the antibody repertoire generated by the host activates antibodies, which can induce immune cell activation and amplify inflammation with subsequent disease severity. We did not monitor the biological function of the antibodies in our patient population. Therefore, we cannot draw conclusions about the presence of neutralizing antibodies in our patient cohort. However, given the finding that the final levels of antibodies were similar in the mild and moderate/severe groups, we believe that the increased morbidity of the moderate/severe patient cohort is not due to differences in antibody function. In support of this notion, it was recently suggested that adaptive immunity toward SARS-CoV-2 resulted from both humoral and cellular immunity^20,21^. In fact, CD4^+^ and CD8^+^ T-cell responses were effectively generated toward SARS-CoV-2 antigens and were correlated with anti-SARS-CoV-2 IgG and IgA titers.

Serological testing will serve in the near future as a powerful tool to conduct epidemiological studies in distinct populations and continents^7^. In addition to such studies, better understating the kinetics of anti-SARS-CoV-2 antibody responses will be critical in various therapeutic settings including utilizing antibodies as part of plasma/antibody therapy or alternatively monitoring the immune response in developing future vaccines. Although we neither monitored the biological function of the antibodies nor the presence of long-lasting memory cells, our study demonstrates a clear reduction in antibody levels 42 days post symptoms. Although this was most prominent in IgA and IgM, a trend toward reduction was also observed in IgG. This is specifically important since vaccines are largely based on generating long-lasting immunity and neutralizing IgG antibodies. Indeed, a recent publication revealed reduced antibody levels (total and neutralization) between the acute phase and the convalescence phase^22^. If, following vaccination, a similar reduction in IgG antibodies will be observed (similar to our findings), the exact vaccination regimen including secondary boosts for the generation of long-lasting memory should be considered.

In summary, our study provides a comprehensive analysis of the antibody response, with specific emphasis on the kinetics of all three major antibody classes toward SARS-CoV-2 RBD and NP antigens in mild and moderate/severe patients. By establishing a rapid, accurate, and robust method as well as analysis, our data have direct methodological implications for future basic research and epidemiological surveys. In addition, our kinetic analysis provides important insights and considerations of future vaccination strategies

## Material and Methods

### Reagents

Unless stated otherwise, all reagents were purchased from Biological Industries, Beit-Haemek, Israel. The SARS-CoV-2 receptor binding domain (RBD) antigen was homemade or purchased from Sigma-Aldrich. SARS-CoV-2 nucleocapsid protein (NP) antigen was purchased from Aalto Bio Reagents (code CK 6404-b). HRP-conjugated secondary antibodies were purchased from Jackson ImmunoResearch Labs (West Grove, PA, USA). Regular ELISA plates were purchased from Greiner Bio-One. All reagents for the electrochemiluminescence test were purchased from MesoScale Diagnostic LLC: (MSD) MULTI-ARRAY® 96 Plate Pack (Cat #: L15XA); Human/NHP IgG Detection Antibody Product (100 ug) (Cat #: D20JL); Human/NHP 1gM Detection Antibody Product (100 ug) (Cat # D20JP); Human/NHP 1gA Detection Antibody Product (100 ug) (Cat # D20JJ); MSD GOLD Read Buffer A (Cat #: R92TG); MSD Blocker A Kit (Cat #: R93AA).

### Expression of recombinant SARS-CoV-2 RBD

The codon optimized sequence of SARS-CoV-2 RBD protein was synthesized by Syntezza-Israel and cloned into the pcDNA 3.1 mammalian expression vector. A hexa-histidine tag (his-tag) was added at the N-terminal for downstream protein purification. The construct was used to transiently transfect Expi293F cells (Thermo Fisher Scientific, Inc.) using the ExpiFectamine 293 Transfection Kit (Thermo Fisher Scientific, Inc.). Seven days post-transfection, the cell supernatant was collected, filtered (0.22µm), and the protein was purified using Ni-NTA (GE Healthcare Life Sciences) affinity chromatography, washed and eluted using 250mM imidazole. The RBD protein was buffer exchanged to PBS, aliquoted, and stored at −80°C.

### Patients and their sample collection

Patients’ samples were obtained from symptomatic individuals testing positive for SARS-Cov-2 by quantitative PCR. Samples were obtained from patients hospitalized at Hasharon Hospital, which is a designated Corona Hospital in Israel. Peripheral blood was obtained (∼5ml) from each patient at different time points including during admission, hospitalization, dismissal, and/or during a routine check-up in the clinic for COVID-19 recovered patients. Samples were also obtained from blood bank donors; they were collected before November 2019. All experiments were reviewed and approved by the Helsinki committee (IRB#RMC-0265-20) and were performed according to their regulations and guidelines.

### Disease severity definition

COVID-19 patients’ disease severity was defined for confirmed COVID-19 patients according to the Israel Ministry of Health as follows:

1. Mild disease: Respiratory disease in the upper airways or pneumonia that does not follow the stated definitions for moderate/severe disease.
2. Moderate disease: Pneumonia with one of the following characterizations (that does not follow the these severe disease definitions):
  a. More than 30 breaths per minute (RR>30/min).
  b. Respiratory distress.
  c. Less than 90% O_2_ saturation in room air.
3. Severe disease: Pneumonia with a respiratory distress of RR>30/min, blood oxygen saturation <90%, respiratory failure [Acute respiratory distress syndrome (ARDS)], sepsis or shock.

### Serum preparation

Whole blood was centrifuged (500xg, 5 minutes) in secure buckets. Supernatant was transferred into a clean 1.7/2ml Eppendorf tube. Thereafter, the serum was inactivated by heat (at 56°C, for 30 minutes). The samples were apportioned into 50μl aliquots and stored at –20°C or –80°C.

### TauMed ELISA protocol

Designated electrochemiluminescence plates were coated with 30μl of purified antigen (RBD or NP at 2μg/ml in PBS) and incubated overnight at 4°C.Thereafter, the plates were washed three times (200μl per well) with MSD washing buffer and blocked with 150μl of MSD blocking buffer per well [1 hour at Room Temp (RT)]. Blocking buffer was tapped out before adding 50μl of the diluted sample to each well [3.6μl of the sample was added to 180μl of the sample diluent (PBST + 1% MSD blocker A)] and incubated for 30 min at RT. Subsequently, the plates were washed three times (200μl per well) and 50μl of detection antibodies were added and incubated for 20 minutes at RT. All of the detection antibodies were diluted in PBST + 1 %MSD blocker A as follows:

IgG detection antibodies to 0.25ug/ml (1:2000)

IgM detection antibodies to 0.25ug/ml (1:2000)

IgA detection antibodies to 0.5ug/ml (1:1000)

Finally, the plates were washed three times (200μl per well) with MSD washing buffer, and 150μl MSD Gold Read buffer was added to each well (avoiding air bubbles in each well). Plates were read within 20 minutes, using MESO QuickPlex SQ 120.

### HRP ELISA

HRP ELISA was performed similar to the above-mentioned protocol with the following changes: 1) blocking was conducted using 3% skim milk; 2) the sample incubation time was 2 hours; 3) the detection antibody incubation time was 1 hour; and 4) HRP substrate was added for 10 minutes and the reaction was stopped using 50μl 1N HCl. Readouts at 405nm (using 595nm background subtraction) were performed within 5 minutes using BioTek EPOCH2.

### Statistical analysis

All of the statistical analyses were performed using GraphPad Prism 8 software. Sensitivity and specificity were determined using ROC analysis (Wilson/Brown 95% CI). To compare ranks, a Nonparametric Mann-Whitney t-test was performed. In comparative assays, a one-way ANOVA nonparametric Kruskal-Wells test, followed by Dunn’s multiple comparisons test, was performed. In all experiments p values <0.05 were considered significant.

## Data Availability

All data will be available upon request

## Author contributions

AM, EBL, IM, GM, GM performed experiments; AM, UQ, YW, DD, MD, MK, CD, SM, GM designed the experiments; TKR, DD, MD, MM, LS, GT, MK, CD, SM, FN, drew blood and provided patient charactaristics; AM and GM analyzed the data; AM, NF, TKR, DC, WY and GM wrote and edited the manuscript; AM and GM supervised the work.

## Supplumentary File

### Supplemental Figures

**Supplementaal Figure 1.**
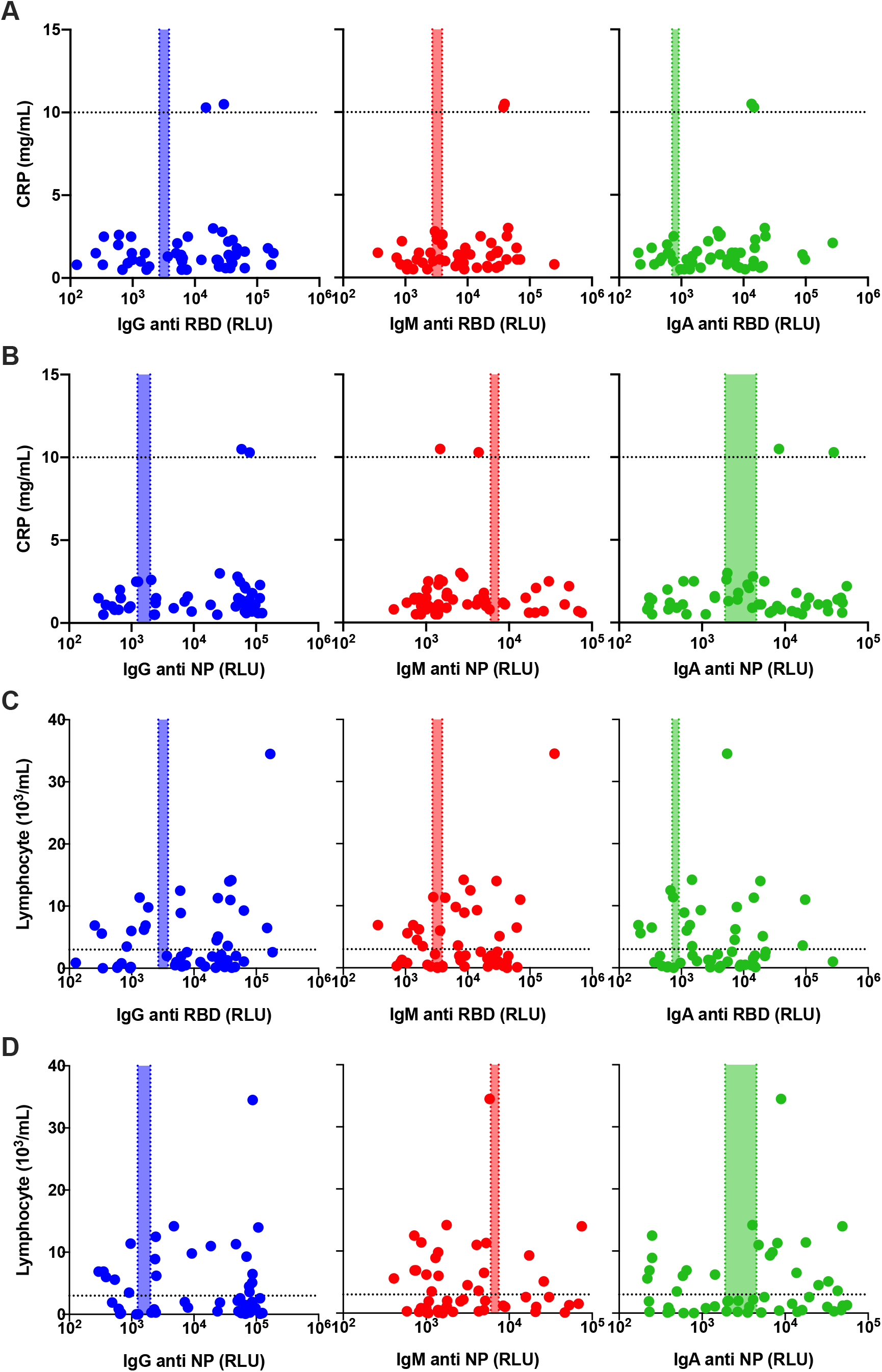
No correlation of antibodies’ response to Lymphocyte count nor to the CRP levels. Peripheral blood was collected from COVID-19 patients. Lymphocyte count and CRP levels were determined immediately. Plasma was obtained, diluted 1:50, and added to a 96-well plate precoated with SARS-CoV-2 RBD (A, C) or NP (B, D) antigens. Patients’ antibody results were graphed against the CRP levels (A-B) and the lymphocyte count (C-D). Data were calculated using GraphPad Prism 8; the dotted X-line represents the calculated cutoff values (95% and 98% sensitivity) discriminating between positive and negative samples, whereas the dotted Y-line represents the cutoff of high levels of either CRP (>10mg/mL) or the lymphocyte count (>3×10^3^/mL). Correlation analysis was performed using a nonparametric Spearman’s correlation test (two-tailed, 95% confidence). No correlation was found.

## References

1. World Health Organization (WHO). Novel Coronavirus (2019-nCoV) Situation Report - 1 21 January 2020. WHO Bulletin (2020).

2. Li, Q. et al.. Early transmission dynamics in Wuhan, China, of novel coronavirus-infected pneumonia. New England Journal of Medicine (2020). doi:10.1056/NEJMoa2001316

3. World Health Organization. Novel Coronavirus (COVID-19) Situation. WHO (June 11) (2020).

4. Remuzzi, A. & Remuzzi, G. COVID-19 and Italy: what next? The Lancet (2020). doi:10.1016/S0140-6736(20)30627-9

5. McKibbin, W. J. & Fernando, R. The Global Macroeconomic Impacts of COVID-19: Seven Scenarios. SSRN Electron. J. (2020). doi:10.2139/ssrn.3547729

6. Li, Y. et al.. Stability issues of RT-PCR testing of SARS-CoV-2 for hospitalized patients clinically diagnosed with COVID-19. J. Med. Virol. (2020). doi:10.1002/jmv.25786

7. Bryant, J. E. et al.. Serology for SARS-CoV-2: Apprehensions, opportunities, and the path forward. Sci. Immunol. (2020). doi:10.1126/sciimmunol.abc6347

8. Stowell, S. & Guarner, J. Role of serology in the COVID-19 pandemic. Clin. Infect. Dis. (2020). doi:10.1093/cid/ciaa510

9. Tay, M. Z., Poh, C. M., Rénia, L., MacAry, P. A. & Ng, L. F. P. The trinity of COVID-19: immunity, inflammation and intervention. Nature Reviews Immunology (2020). doi:10.1038/s41577-020-0311-8

10. Zhao, Z. et al.. A multiplex assay combining insulin, GAD, IA-2 and transglutaminase autoantibodies to facilitate screening for pre-type 1 diabetes and celiac disease. J. Immunol. Methods (2016). doi:10.1016/j.jim.2016.01.011

11. Dabitao, D., Margolick, J. B., Lopez, J. & Bream, J. H. Multiplex measurement of proinflammatory cytokines in human serum: Comparison of the Meso Scale Discovery electrochemiluminescence assay and the Cytometric Bead Array. J. Immunol. Methods (2011). doi:10.1016/j.jim.2011.06.033

12. Amanna, I. J., Carlson, N. E. & Slifka, M. K. Duration of humoral immunity to common viral and vaccine antigens. N. Engl. J. Med. (2007). doi:10.1056/NEJMoa066092

13. Jin, J. M. et al.. Gender Differences in Patients With COVID-19: Focus on Severity and Mortality. Front. Public Heal. (2020). doi:10.3389/fpubh.2020.00152

14. Krammer, F. & Simon, V. Serology assays to manage COVID-19. Science (2020). doi:10.1126/science.abc1227

15. Long, Q. X. et al.. Antibody responses to SARS-CoV-2 in patients with COVID-19. Nat. Med. (2020). doi:10.1038/s41591-020-0897-1

16. Yong, S. E. F. et al.. Connecting clusters of COVID-19: an epidemiological and serological investigation. Lancet. Infect. Dis. (2020). doi:10.1016/S1473-3099(20)30273-5

17. Yongchen, Z. et al.. Different longitudinal patterns of nucleic acid and serology testing results based on disease severity of COVID-19 patients. Emerging microbes & infections (2020). doi:10.1080/22221751.2020.1756699

18. Pedersen, S. F. & Ho, Y.-C. SARS-CoV-2: A Storm is Raging. J. Clin. Invest. (2020). doi:10.1172/jci137647

19. Tetro, J. A. Is COVID-19 receiving ADE from other coronaviruses? Microbes Infect. (2020). doi:10.1016/j.micinf.2020.02.006

20. Ni, L. et al.. Detection of SARS-CoV-2-Specific Humoral and Cellular Immunity in COVID-19 Convalescent Individuals. Immunity (2020). doi:10.1016/j.immuni.2020.04.023

21. Grifoni, A. et al.. Targets of T Cell Responses to SARS-CoV-2 Coronavirus in Humans with COVID-19 Disease and Unexposed Individuals. Cell (2020). doi:10.1016/j.cell.2020.05.015

22. Long, Q.-X. et al.. Clinical and immunological assessment of asymptomatic SARS-CoV-2 infections. Nat. Med. (2020). doi:10.1038/s41591-020-0965-6

